# Genomic Analysis of *Salmonella* Typhi from a Typhoid Conjugate Vaccine Trial

**DOI:** 10.64898/2026.01.06.25342561

**Authors:** Belson M. Kutambe, Priyanka D. Patel, Kenneth Chizani, Niza Silungwe, Christopher Kukacha, Megan E. Carey, James E. Meiring, Matthew B. Laurens, Melita A. Gordon, Philip M. Ashton

## Abstract

**Background:** *Salmonella enterica* serovar Typhi (*S.* Typhi), the causative agent of typhoid fever, continues to pose a major public health challenge in many low- and middle-income countries (LMICs). The World Health Organization recommended typhoid conjugate vaccine (TCV) use in countries with a high burden of disease, and/or high rates of antimicrobial resistance (AMR). Recent introductions of TCVs into national immunisation programmes are expected to substantially reduce this burden. However, the impact of vaccine introduction on pathogen populations has not been widely investigated.

**Methods:** To fill this research gap, we analysed the genomes of isolates from a trial in Malawi. A Phase 3, double-blind, randomized, controlled trial enrolled 28,130 healthy children aged 9 months through 12 years of age to receive either TCV (Typbar-TCV, Bharat Biotech) or meningococcal A conjugate vaccine (MenA). We conducted Illumina whole genome sequencing and compared isolates from the TCV intervention arm to the MenA control arm with regard to: (i) *S.* Typhi lineage distribution, (ii) AMR profile, (iii) mutations in genes associated with Vi capsule biosynthesis and expression, and (iv) phylogenetic population structure.

**Results:** We obtained high-quality genome sequences for 136 *S.* Typhi isolates (24 from the TCV arm, 112 from the MenA arm). Of these, 135 (99%) belonged to lineage L4.3.1.1.EA1. Isolates from the two arms were intermixed across multiple sub-clades. Nearly all isolates (135/136) carried genes encoding resistance to ampicillin, cotrimoxazole, and chloramphenicol. Non-synonymous mutations in the quinolone resistance determining region were identified in five isolates (three GyrA S83F; two GyrA S464F) and no statistically significant difference in prevalence of these mutations was observed between study arms (Fisher’s exact test, *P* = 0.25). Mutations in Vi capsule–associated genes were detected in 11/112 (9%) MenA isolates and 1/24 (4%) TCV isolates (Fisher’s exact test, *P* = 0.67). 12 MenA participants were hospitalised, one of whom had Vi-associated mutations associated with increased virulence in a murine model; however, there was no significant association between tested Vi mutations and hospitalisation (*P* > 0.99).

**Conclusions:** We identified no clinically meaningful genomic differences between *S.* Typhi isolates from participants vaccinated with TCV or MenA, and no Vi-negative *S.* Typhi were detected. These findings suggest no detectable short-term evolutionary impact of TCV on circulating *S.* Typhi populations in this Phase 3 trial. However, given the limited follow-up period and *S.* Typhi’s relatively slow substitution rate, continued genomic surveillance in this population after national TCV introduction in 2023 is essential to detect evolutionary responses to vaccine pressure.

## Introduction

Typhoid fever, caused by *Salmonella enterica* serovar Typhi (*S*. Typhi), remains a significant public health challenge, particularly in low-resource settings [1]. In recent decades, the emergence and spread of several multidrug-resistant (MDR [i.e., resistant to the first-line agents chloramphenicol, ampicillin, and cotrimoxazole]) *S.* Typhi lineages, particularly H58 (clade 4.3.1) and H56 (clade 3.1.1) has complicated antimicrobial treatment and increased the urgency for effective prevention strategies [2]. Furthermore, *S.* Typhi can gain resistance to fluoroquinolones [2,3], third generation cephalosporins [4], and azithromycin [5]. Typhoid conjugate vaccines (TCVs) represent a key public health tool to control typhoid fever and associated antimicrobial resistance (AMR), both by preventing drug-resistant *S*. Typhi infections and potentially by reducing antimicrobial use [6]. These highly effective vaccines target the Vi-capsule and have been recently introduced in some high burden countries [7].

Monitoring pathogen population response to vaccine pressure is crucial to detect potential vaccine escape or shifts in resistance profiles. Previous studies on circulating *Streptococcus pneumoniae* organisms have demonstrated the potential response of pathogen populations to vaccine introduction [8–11]. Further, *S.* Typhi isolates lacking the Vi antigen have been reported as causing disease in Pakistan [12]. In this study, we aimed to provide high-resolution insight into the short-term evolutionary dynamics of *S.* Typhi following TCV use by sequencing isolates collected during a Phase 3 vaccine trial in Malawi. We hypothesized that widespread TCV may exert a selective pressure on *S.* Typhi populations potentially increasing the frequency of mutations in the Vi encoding genes within the TCV group than the MenA group. Data collected during trial passive surveillance follow-up demonstrated that TCV use has resulted in fewer antibiotic prescriptions [13]; hence, we hypothesized that lower transmission of *S.* Typhi and decreased selection pressure due to decreased antimicrobial use might result in lower frequency of AMR genes in TCV recipients.

## Methods

### Sample Collection and Isolation

As part of a previously published vaccine trial, 28,130 children were vaccinated with either TCV (n = 14,069) or MenA (n = 14,061) [7,14]. If trial participants presented with febrile illness (subjective fever for ≥72 hours, an axillary temperature of ≥38°C, or hospitalization with a history of fever of any duration), a blood culture was obtained (5 ml in children <5 years of age or 10 ml in children ≥5 years of age). AMR profiles of *S.* Typhi isolates were tested using disk diffusion. Isolates exhibiting pefloxacin resistance underwent confirmatory testing for ciprofloxacin resistance using E-test (bioMérieux), with a minimum inhibitory concentration of more than 0.06 mg per litre indicating non-susceptibility. Full details of study procedures can be found in Patel et al., 2021 [7,14].

### Microbiology and Molecular Biology

Before sequencing, isolates were cultured overnight on XLD agar at 37°C and genomic DNA was extracted with the Wizard Genomic DNA Extraction Kit (Promega, Madison, WI, USA) following the manufacturer’s recommendations. The first batch of genomic DNA was shipped to the Wellcome Sanger Institute for indexed whole genome sequencing on an Illumina HiSeq 2500 or HiSeq 4000 platform (Illumina, San Diego, CA, USA) to generate paired-end reads of 100 bp in length.

### Bioinformatics

Raw reads were processed using the Bactopia pipeline v.3.1.0 [15], which included quality control (https://github.com/s-andrews/FastQC), read trimming, variant calling (https://github.com/tseemann/snippy), and lineage assignment (ߢbactopia --samples samples.txt -profile docker --outdir bactopia --use_bakta true --bakta_db /path/to/bakta_db – ask_merlin tru’). We used the CT18 reference strain of *S*. Typhi ( https://www.ncbi.nlm.nih.gov/nuccore/AL513382.1) as the reference for variant calling via the PHEnix pipeline (https://github.com/ukhsa-collaboration/PHEnix). Resulting consensus sequences were masked for highly variable and repetitive regions [16] using Bedtools v.2.3.0 [17]. A maximum likelihood phylogenetic tree was inferred using IQ-TREE v2.3.5 [18] with in-built model selection (TVM+F+I,best fit model chosen according to BIC), and tree visualization was performed using iTOL v6.8 [19]. To place our sequencing results into wider temporal context, we included in the tree the previously sequenced *S.* Typhi isolated from Blantyre (n=302) that were not part of the Phase 3 trial (herein referred to as non-trial Blantyre *S.* Typhi isolates). Genes in the viaA regulatory region (represented by *rcsC*), the viaB locus (*tviA*–*tviE* and *vexA*–*vexE*), and the OmpB locus (*ompR* and *envZ*), which together regulate Vi capsular polysaccharide expression, were included in the analysis [20–22] (Figure 1).

**Figure 1:**
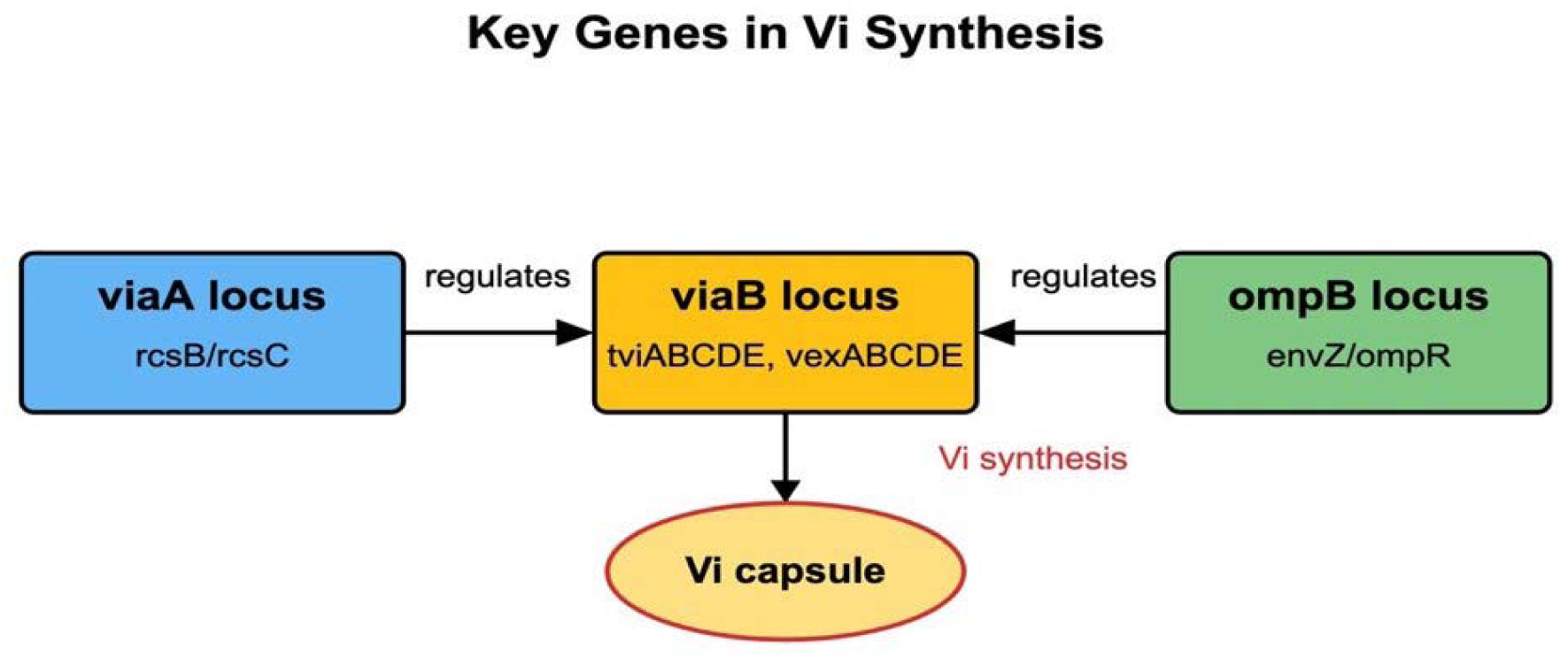
Key genes involved in Vi capsule synthesis included in the analysis.

To investigate whether H58 isolates contained the IncH1 plasmid that commonly carries the MDR gene cassette associated with H58 *S*. Typhi organisms, PlasmidFinder (Danish Technical University, Denmark: http://cge.cbs.dtu.dk/services/PlasmidFinder/) was used to detect plasmid presence.

### Statistical analysis

Two-by-two contingency tables were constructed to assess: (i) the association between the proportion of isolates with QRDR mutations and vaccine arm; (ii) the association between the proportion of isolates with Vi-capsule mutations and vaccine arm; and (iii) the association between the presence of virulence-modifying SNPs and hospitalization among MenA participants. These analyses were performed using the OpenEpi two-by-two module (https://www.openepi.com/TwobyTwo/TwobyTwo.htm).

R version 4.4.1 was used to visualize the distribution of Vi mutations across vaccine arms and to display the association between virulence-modifying SNPs and hospitalization among MenA participants.

### Ethical approval

Ethics approval was obtained from the Malawi National Health and Sciences Research Committee, and from the University of Liverpool Ethics Review Board and the University of Maryland Baltimore Institutional Review Board. The sponsors had no role in the design of the trial; the collection, analysis, or interpretation of the data; or the writing of the manuscript.

### Data summary

The authors confirm all supporting data, code and protocols have been provided within the article or through supplementary data file. The raw sequencing reads for the 136 *S*. Typhi isolates subjected to whole genome sequencing have been deposited in the European Nucleotide Archive (ENA) under Bioproject PRJEB104623 (see supplementary data for details).

## Results

A total of 136 isolates were sequenced from 134 participants (Supplementary Table 1). Two participants, one from TCV and one from MenA, each contributed two isolates originating from distinct clinical episodes. Of these, 135/136 isolates (24 from the TCV arm and 111 from the MenA arm) belong to the L4.3.1.1.EA1 genotype, and the remaining isolate (from the MenA arm) was assigned to lineage 2.2. Importantly, no Vi-negative *S.* Typhi were detected in the trial. In a maximum likelihood phylogenetic tree, no clustering by vaccine arm was observed (Figure 2). Placing our sequencing results within a broader temporal context shows that H58 (lineage L4.3.1.1.EA1) continues to dominate the *S. Typhi* population in Blantyre (76%, 334/438), with limited genetic diversity observed between the Phase 3 trial isolates and the non-trial Blantyre isolates. PlasmidFinder results suggested that the IncH1 plasmid was not present in any H58 isolate.

**Figure 2:**
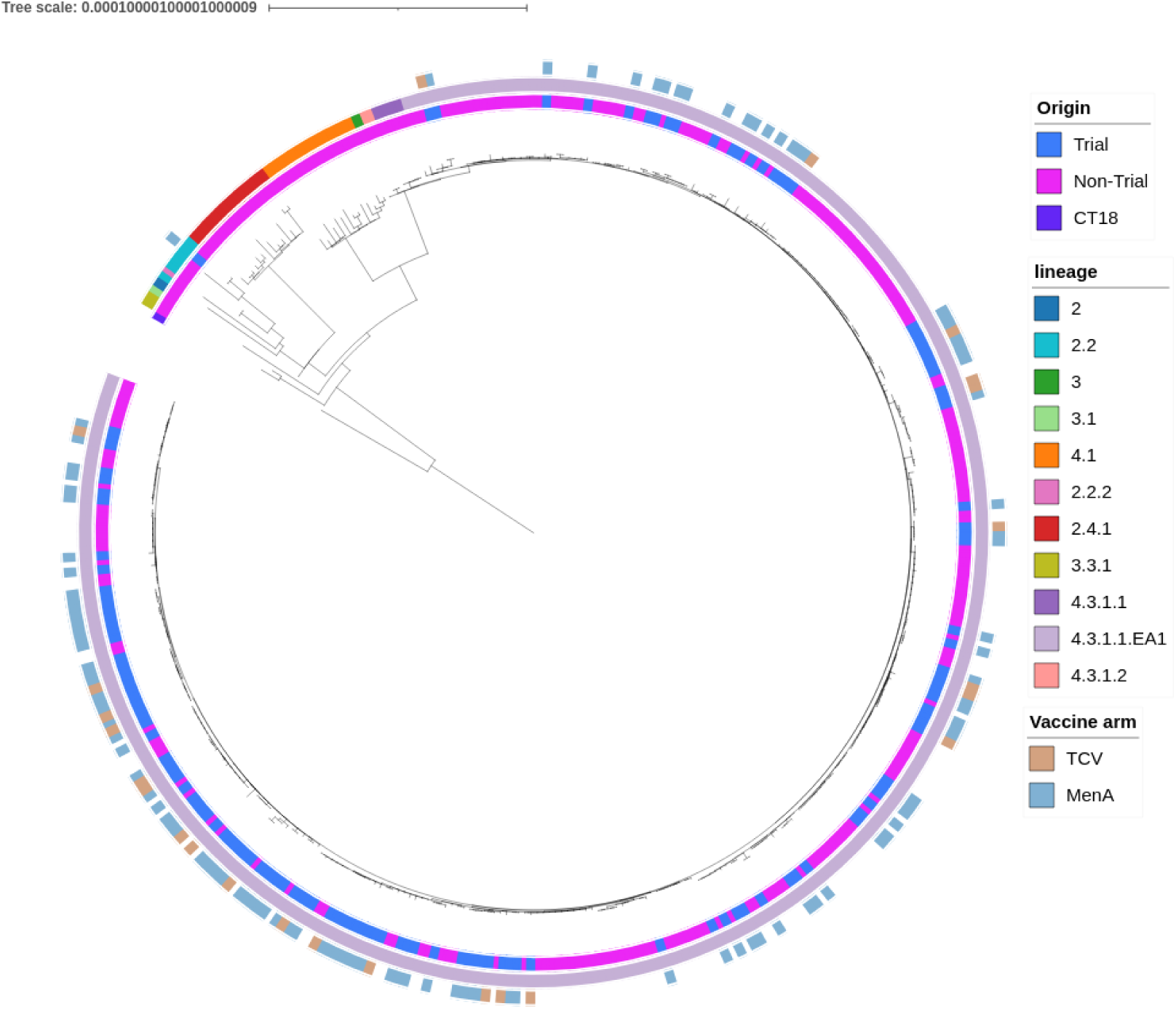
Maximum-likelihood phylogeny of 136 Phase 3 trial isolates contextualized with 302 previously sequenced *S. Typhi* isolates from Blantyre (innermost ring) The middle ring depicts the overall lineage distribution for both the trial and non-trial isolates. The outer ring shows the lineage distribution stratified by vaccine arm for the trial isolates.

### Antimicrobial Resistance (AMR) characterization

Nearly all the trial *S.* Typhi isolates sequenced (99%, 135/136) encode genes associated with an MDR phenotype *aph(6)-Id,aph(3’’)-Ib, blaTEM-1*, *dfrA7*, *catA1*, *sul1*, and *sul2*. The MDR profile distribution was the same across both TCV and MenA arms, with no differences observed (Fisher’s exact test P > 0.99). Five isolates (3%, 5/136) with quinolone resistance determining region (QRDR) mutations were observed; three isolates encoded GyrA S83F and two encoded GyrA S464F. Of the isolates with QRDR mutations, three were from the MenA arm and two were from the TCV arm (Fisher’s exact test P = 0.25, Supplementary Table 3).

### Vi Locus Variants

The *S.* Typhi genes encoding the Vi biosynthesis system were present (>= 99% of bases covered at >= 5x depth) in all isolates. 12 non-synonymous SNPs in the Vi-capsule encoding genes were identified in 11 of the 136 isolates (Figure 3, Supplementary Table 2). Of the 11 isolates, 10 were from the MenA arm, and one from the TCV arm (Figure 3). Trial arm was not statistically associated with detection of SNPs in Vi capsule encoding genes (Fisher’s exact test p=0.67, Supplementary Table 3). No indels were detected. The SNPs were distributed among the following genes: *tviD* (locus tag STY4659; *n* = 6), *tviE* (locus tag STY4656; *n* = 5), and *vexA* (locus tag STY4655; *n* = 1).

**Figure 3:**
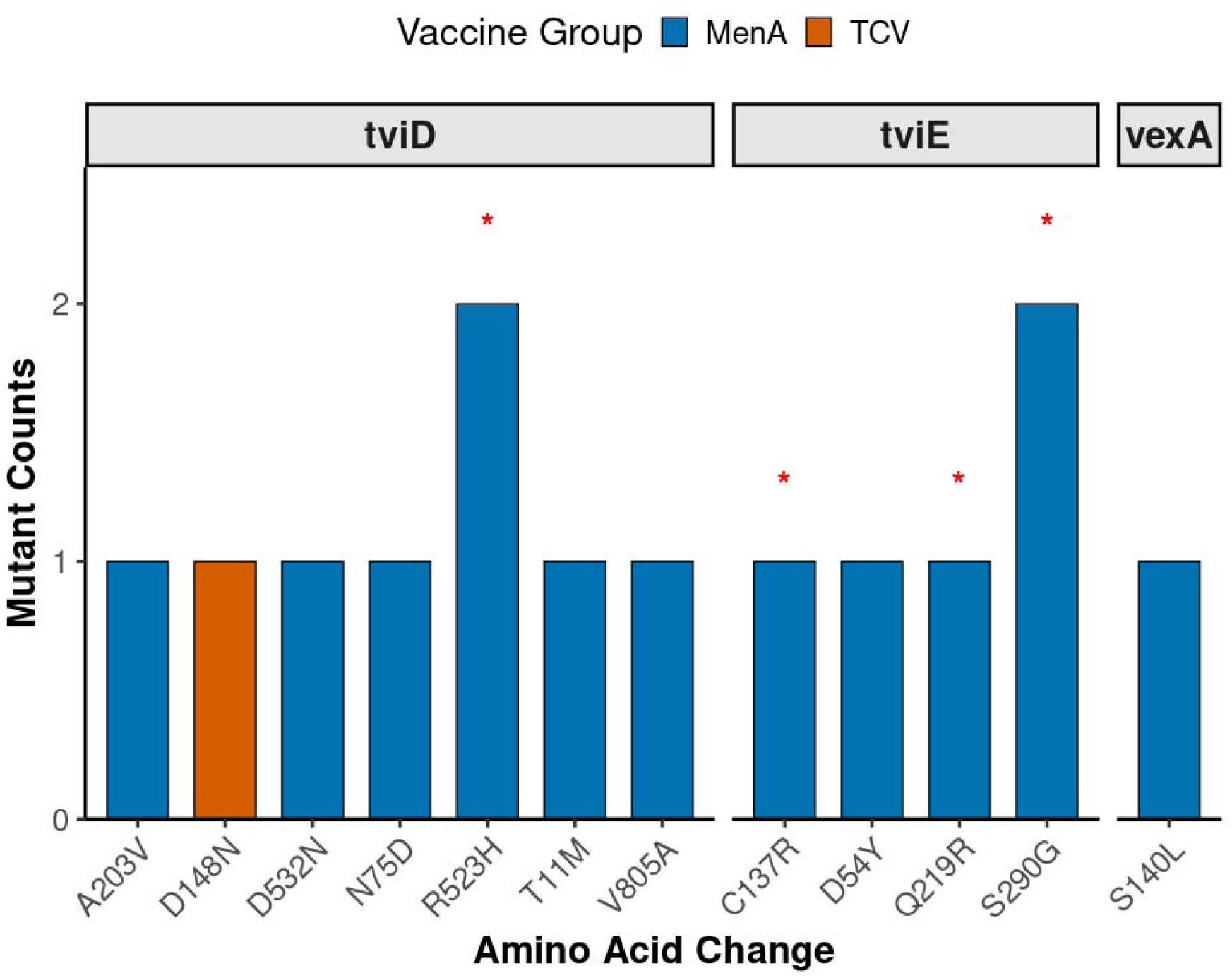
Distribution of Vi antigen mutations among trial isolates. Amino acid changes previously reported to confer hyper-virulence in murine models are marked with an asterisk (*). **TCV** refers to Vi mutations identified in isolates from participants who received the Typhoid Conjugate Vaccine, while **MenA** refers to Vi mutations observed in isolates from participants who received the meningococcal A conjugate vaccine.

Four SNPs encoding amino acid changes R523H in the TviD protein and Q219R, S290G, and C137R in the TviE protein have previously been reported to increase the virulence of *S.* Typhi infections in mice [20] (Figure 4). As these SNPs were also identified in our dataset, we analysed their potential association with disease severity in humans, using hospitalization as a proxy for severity (Figure 4). This analysis was restricted to 112 MenA arm isolates, as the effect of TCV on disease severity in breakthrough infections remains unclear. Of the 111 participants with sequenced *S.* Typhi, 4 were hospitalized. One hospitalized patient was infected with an isolate carrying Vi-capsule SNPs (TviD A203V and TviE S290G), and the three other hospitalized patients had no Vi-capsule SNPs; Vi capsule SNPs were not associated with having previously reported virulence increasing Vi mutation and hospitalization (Fisher exact test p > 0.99, Supplementary Table 3).

**Figure 4:**
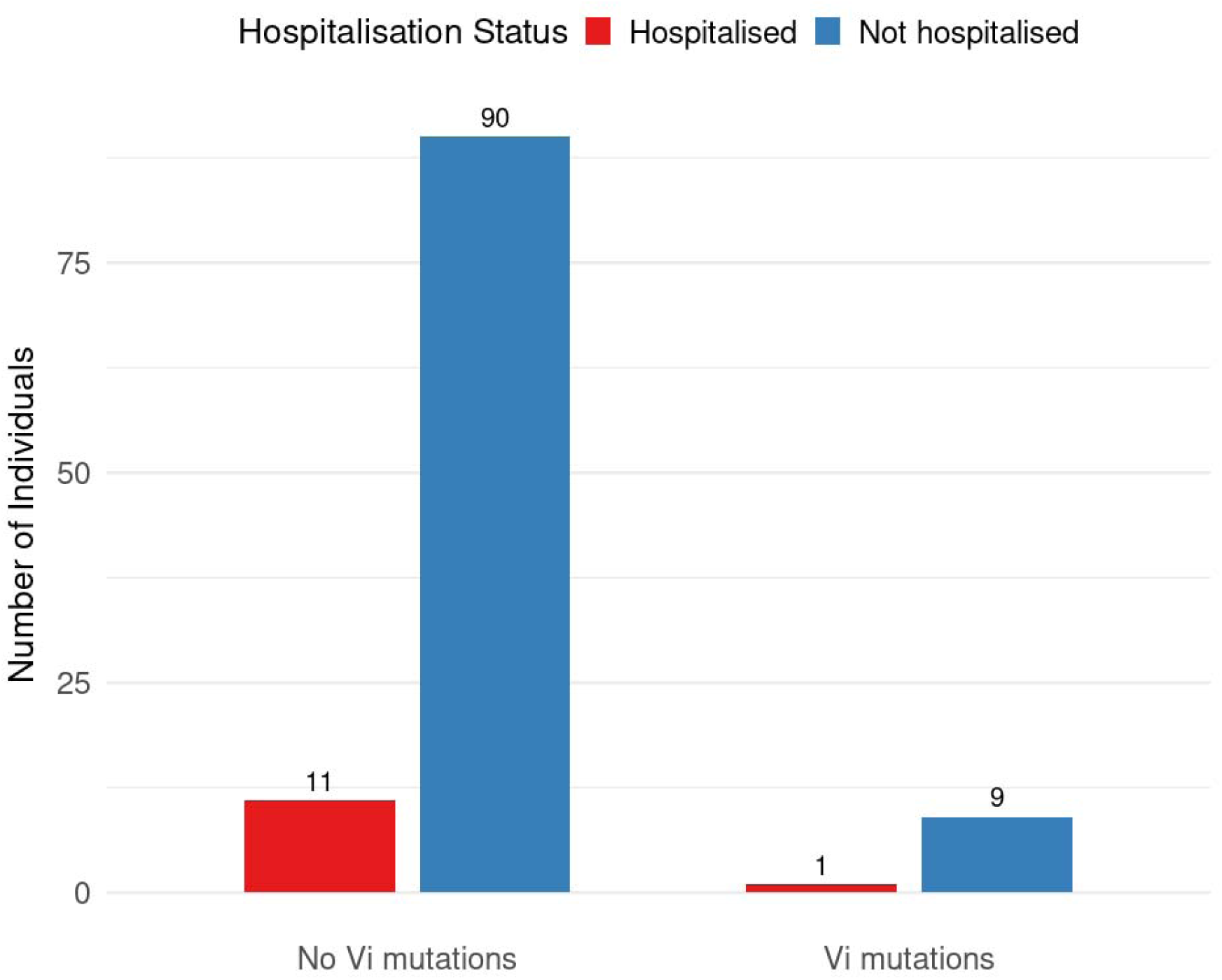
Association between previously reported virulence-enhancing Vi-capsule SNPs and hospitalization among MenA participants. Fisher’s exact test: p > 0.99.

### Non-Typhoidal Salmonella (NTS)

One potential consequence of introducing a Vi-based vaccine is the emergence of Vi-negative S. Typhi, which could be misclassified in our laboratory workflow as non-typhoidal Salmonella (NTS). To exclude this possibility, we examined all NTS isolates recovered during the trial. A total of 20 NTS isolates were obtained from children enrolled in the Phase 3 trial. High quality sequencing data were generated for 17 of these isolates; 9 from the MenA arm and 8 from the TCV arm. 16/17 of the isolates were *Salmonella enterica* serovar Typhimurium (*S.* Typhimurium), and one was *Salmonella* Bovismorbificans. Among the *S.* Typhimurium isolates,12 were ST313, two were ST19 and two were ST3257, which is a single locus variant of ST313. Overall, the following AMR genes were found at most in these NTS isolates; *aadA1*, *aph(3’’)-Ib*, *aph(6)-Id*, *blaTEM-1*, *catA1*, *dfrA1*, *sul1,* and *sul2*.

## Discussion

No consistent genetic differences were identified between *S.* Typhi isolates from the TCV and control (MenA) arms of the Phase 3 vaccine trial. There is no phylogenetic clustering by vaccine arm, no differences in AMR or genotype distribution between vaccine arms, no differences in frequency of Vi mutations between vaccine arms, and no Vi-negative *S.* Typhi were detected.

The population structure and AMR patterns of *S*. Typhi isolates during the vaccine trial reveal that MDR L4.3.1.1.EA1 sub-lineage dominates the circulating *S*. Typhi population in Blantyre. This is consistent with previous reports describing the dominance of MDR H58 lineages in Blantyre and globally [23,24]. Our analysis shows no evidence of an association between sub-clade membership within L.4.3.1.1.EA1 and vaccine arm; isolates from vaccinated and unvaccinated participants were distributed across the same sub-clades.

We hypothesized that the reduced antimicrobial use observed in the TCV arm of the trial [13] might translate into fewer AMR genes among *S*. Typhi isolated from TCV recipients. However, this was not supported by our findings. Two factors likely explain this: (1) the dominance of MDR H58 across both vaccine groups, and (2) because AMR genes in Blantyre H58 isolates are chromosomally integrated, the MDR phenotype is expected to be stable regardless of immediate antibiotic pressure. [23]. The lack of a reduction in AMR genes in *S.* Typhi could also be related to the short follow-up. Indeed, our plasmid analysis revealed the absence of the IncHI1 plasmid, which typically carries MDR regions in H58, confirming chromosomal integration. This finding implies that even in the context of reduced antibiotic pressure, MDR genes may persist in H58 due to stable genomic integration.

Our primary hypothesis was that TCV pressure might drive the emergence of Vi antigen variants capable of evading TCV-induced immunity. Although amino acid altering Vi substitutions were identified, the distribution was similar between the intervention and control arms, which supports the null hypothesis. Our analysis identified no association between the four amino acid changes (R523H, Q219R, S290G, and C137R) located in the TviD and TviE proteins, previously shown to increase virulence in murine models [20] and disease severity using hospitalization as a proxy. A limitation to both these findings is the small number of isolates from the TCV arm, and the relatively small number of hospitalisations, which limited statistical power.

One way in which vaccine escape could manifest is in Vi-negative *S.* Typhi [12] which would be typed as non-typhoidal Salmonella in our laboratory workflow. In addition, 20 non-typhoidal *Salmonella* (NTS) isolates were recovered from blood cultures, all genomically confirmed as NTS. The detection of NTS during the trial validates the laboratory serotyping of the trial isolates and confirms the absence of Vi-negative *S*. Typhi in circulation.

Our study has several strengths that enhance its significance. First, it was nested within a clinical trial with good coverage of the study population. Second, genome sequencing of the isolates enabled investigation of multiple genetic aspects, including phylogenetic relationships, AMR determinants, and specific SNPs within the Vi biosynthesis region. Finally, the study allowed us to evaluate mutations previously associated with virulence in mouse models within a human clinical context.

However, our study also has limitations. The small number of isolates, especially from vaccinees, limited statistical power to detect subtle vaccine-related genetic differences. Since this study was nested within a clinical trial, we had a relatively brief period of follow up. The low genetic diversity of *S.* Typhi circulating in Blantyre also limits the power to detect population changes in terms of lineage distribution. Additionally, using hospitalization as a proxy for disease severity does not capture the complete clinical spectrum of typhoid infection. Finally, markers of potential hypo- or hypervirulence identified in murine models require validation in larger datasets with clinical metadata and vaccination status.

To our knowledge, there are no peer-reviewed or preprint reports that present whole genome comparisons of *S.* Typhi isolates stratified by vaccine arm from a randomized TCV trial. Most published genomic work addresses population surveillance or pre/post-introduction comparisons rather than a comparison of trial arms within a randomized study. For example, Gaetan *et al.* conducted a genomic epidemiology study of *S.* Typhi in Harare, Zimbabwe (2012–2019)[25], before the national rollout of the TCV, and Silva *et al.* studied the population structure and AMR profiles of 174 *S.* Typhi and 54 *S.* Paratyphi A isolates in the context of a phased municipal vaccination campaign in Navi Mumbai, India[26].

No evidence for vaccine escape mutations in this phase 3 clinical trial setting was detected, but the limitations outlined above mean our findings should be interpreted with caution, and further research with longer follow-up and a larger sample size is required to confirm findings. Given the limited number of participants and follow-up period, continued genomic surveillance in this population after TCV introduction in 2023 will be essential to monitor longer-term evolutionary responses to vaccine pressure in the larger population.

## Conclusion

This study provides an early assessment of *S*. Typhi genomic evolution after TCV introduction in Malawi. Our findings indicate limited divergence between vaccine and control arms, stable AMR profiles, and no significant difference in mutations affecting the phenotype of Vi-capsule genes. Continued monitoring of *S*. Typhi populations is essential to track the long-term impact of vaccination on pathogen evolution.

## Data Availability

The authors confirm all supporting data, code and protocols have been provided within the article or through supplementary data file. The raw sequencing reads for the 136 S. Typhi isolates subjected to whole genome sequencing have been deposited in the European Nucleotide Archive (ENA) under Bioproject PRJEB104623 (see supplementary data for details).

## Author Contribution

BMK: conceptualisation, data curation, formal analysis, investigation, methodology, software, visualisation, writing (original draft), writing (review and editing). PDP: data curation, investigation, writing (review and editing). KC: investigation, writing (review and editing). NS: investigation, writing (review and editing) CK: investigation, writing (review and editing). MEC: investigation, writing (review and editing) JEM: data curation, investigation, writing (review and editing). MBL: funding acquisition, investigation, writing (review and editing). PMA: project administration, supervision, conceptualisation, data curation, investigation, writing (review and editing). MAG: project administration, supervision, conceptualisation, funding acquisition, writing (review and editing). All authors had full access to all the data in the study and accept responsibility for the decision to submit for publication. PMA and MAG verified the data underlying the study.

## Acknowledgements

We thank the trial participants and their parents and guardians; the staff of the Malawi Ministries of Health and Education; the Blantyre District Health Officer; the staff of the Paediatric Department of the Queen Elizabeth Central Hospital; the staff of the health centers in Ndirande and Zingwangwa, the Gateway Health Clinic, and the Nancholi Youth Organization Clinic; the director and the staff of the departments of the Malawi–Liverpool–Wellcome Program; the Malawi trial team (John Ndaferankhande, Nedson Chasweka, Lucky Somanje, Chrissy Banda, Josophine Chilongo, Patricia Phula, Georgina Makuta, Monica Kamwana, and Moses Kamzati); Victoria Mapemba (Blantyre Malaria Project); the University of Maryland team (Leslie Jamka, Shrimati Datta, Ian Woods, Christina Scheele, and Tamar Pair); the members of the data and safety monitoring board (Roma Chilengi [chair], Prakash Ghimire, A.K.M. Nurul Anwar [deceased], S.M. Shamsuzzaman, Jalaluddin Ashraful Haq, Nur Haque Alam, Tisungane Knox Titus Mvalo, and Mary E. Putt); the staff of the Centre for Genomic Research, University of Liverpool; and Bharat Biotech International for supplying the investigational vaccine free of charge.

